# The Social Factors, Epigenomics, and Lupus in African American Women (SELA) study: protocol for an observational mechanistic study examining the interplay of multiple individual and social factors on lupus outcomes in a health disparity population

**DOI:** 10.1101/2022.03.09.22272149

**Authors:** Emily L. Vara, Carl D. Langefeld, Bethany J. Wolf, Timothy D. Howard, Gregory A. Hawkins, Queen Quet, Lee H. Moultrie, L. Quinnette King, Ivan D. Molano, Stephanie L. Bray, Lori Ann Ueberroth, S. Sam Lim, Edith L. Williams, Diane L Kamen, Paula S. Ramos

## Abstract

**Introduction:** Despite the disproportional impact of systemic lupus erythematosus (SLE) on historically marginalized racial and ethnic communities, the individual and sociocultural factors underlying these health disparities remain elusive. We report the design and methods for a study aimed at identifying the epigenetic mechanisms by which risk and resiliency social factors affect gene function and thereby influence SLE in a health disparity population.

**Methods and analysis:** The Social Factors, Epigenomics, and Lupus in African American Women (SELA) study is a cross-sectional, case-control study involving the Medical University of South Carolina, Emory University, and Wake Forest School of Medicine. A total of 600 self-reported African American females will be invited to participate. All participants will respond to questionnaires that capture detailed sociodemographic and medical history, validated measures of racial discrimination, vicarious racism stress, social support, healthcare utilization and lost productivity, as well as disease activity and damage for cases. Physician-reported disease activity will also be incorporated Participants will choose if they wish to receive their genetic ancestry estimates and be involved in research. Blood samples are required to provide serum, plasma, PBMCs counts, DNA and RNA. The primary goals of SELA are to identify variation in DNA methylation (DNAm) associated with self-reported exposure to racial discrimination and exposure to social support, to evaluate whether social DNAm sites affect gene expression, to identify the synergistic effects of social factors on DNAm changes on SLE, and to develop a social factors-DNAm predictive model for disease outcomes. This study was approved by and will be conducted in cooperation with the Sea Island Families Project Citizen Advisory Committee.

**Discussion and dissemination:** SELA will respond to the pressing need to identify the regulatory mechanisms through which social exposures influence SLE in a health disparity population, clarify the interplay and underlying mechanism by which various positive and negative social determinants of health influence epigenomic variation, and how the resulting biological changes may contribute to the lupus health disparity. Results will be published and shared with patients and the community. These findings may inform the development of psychosocial interventions that prevent or mitigate risk exposures, and services or interventions that promote positive exposures. Development of these novel treatments and preventative interventions, as informed by the results of this study, is paramount to the closure of the health disparities gap.

## Introduction

Health disparities in systemic lupus erythematosus (SLE) are well established. As recently reviewed, women are 8-10 times more likely than men to develop SLE; relative to European Americans, African Americans are 3-4 times more likely to develop SLE, suffer from remarkably higher disease severity and death rates, and are more likely to suffer from multiple comorbidities such as depression, cardiovascular disease, diabetes, and worse health-related quality of life.^1^ SLE is among the leading causes of death in young females (highest for African American and Hispanic females),^2 3^ underscoring its impact as an important public health issue.

Despite the disproportionate impact of SLE on minority racial and ethnic communities, the causes for these health disparities remain elusive. The causal mechanisms underlying SLE risk and outcomes among and within ethnic groups are complex, involving biological, sociocultural, physical, and other environmental exposures. Differences in disease risk allele frequency in populations might underlie some of the health disparities, as multiple genetic risk factors for SLE vary among populations.^4^ Additionally, multiple social stressors (e.g., poverty, low household income, unemployment, perceived stress, racial discrimination) negatively affect SLE outcomes, while protective factors (e.g., social support, healthy lifestyles) can help improve SLE outcomes.^5^ However, the mechanisms by which these adverse and protective social factors synergistically modulate disease outcomes are not currently well understood.

Most SLE research to date has focused on biological mechanisms independent of the effects of social exposures, and health disparities research has focused primarily on the influence of socioeconomic determinants on outcomes without considering the biological mechanisms involved. This has resulted in a knowledge gap regarding the interactions among individual and social factors that contribute to disparities in SLE outcomes. We propose a socioecological model of SLE outcomes that emphasizes the importance of integrating sociocultural and individual determinants to understand and address health disparities in SLE.^1^

Despite the influence that social or environmental experiences have on SLE in African American women, it is not known how these experiences influence disease outcomes. We postulate that in African American women, exposure to adverse and protective social contexts is associated with epigenomic changes that in turn are associated with disease outcomes. We specifically hypothesize that social support compensates for the detrimental, independent effect of racism on SLE, considering other sociodemographic and behavioral characteristics, through epigenetic and gene regulatory mechanisms. We will investigate the role of DNA methylation in mediating the effects of social exposures on SLE. DNA methylation levels are associated with psychosocial factors, including socioeconomic status and general perceived stress.^6–8^ Additionally, although DNA methylation varies between populations, and this variation is partially explained by their distinct genetic ancestry, environmental factors not captured by ancestry are significant contributors to variation in DNA methylation, underscoring the notion that an interaction between social, genetic and epigenetic factors underlies the health disparity in SLE.^9–16^

The goal of this study is to identify epigenetic mechanisms by which both positive and negative social factors affect gene function and therefore influence SLE in African American women. To understand the effects of positive and negative social environments on SLE through epigenomic changes, we will test a conceptual model that integrates multiple social, demographic, behavioral, genomic, epigenomic, and transcriptomic factors. The research will identify the epigenetic mechanisms by which risk and resiliency factors affect gene function and thereby influence SLE in African American women.

## Methods and Analysis

### SELA study overview

Social Factors, Epigenomics, and Lupus in African American Women (SELA) is an observational study whose goal is to evaluate the effects of racial discrimination and social support on SLE outcomes through changes in DNAm and gene expression (figure 1). The study has three major aims: (Aim 1) To identify variation in DNA methylation (DNAm) associated with self-reported (a) exposure to racial discrimination, (b) exposure to social support, and (c) assess whether these exposures and SLE are associated with epigenetic age acceleration; (Aim 2) To assess whether social DNAm sites affect gene expression; (Aim 3) To identify the synergistic effects of social factors on DNAm changes on SLE and develop a social factors-DNAm predictive model for disease outcomes.

**Figure 1.**
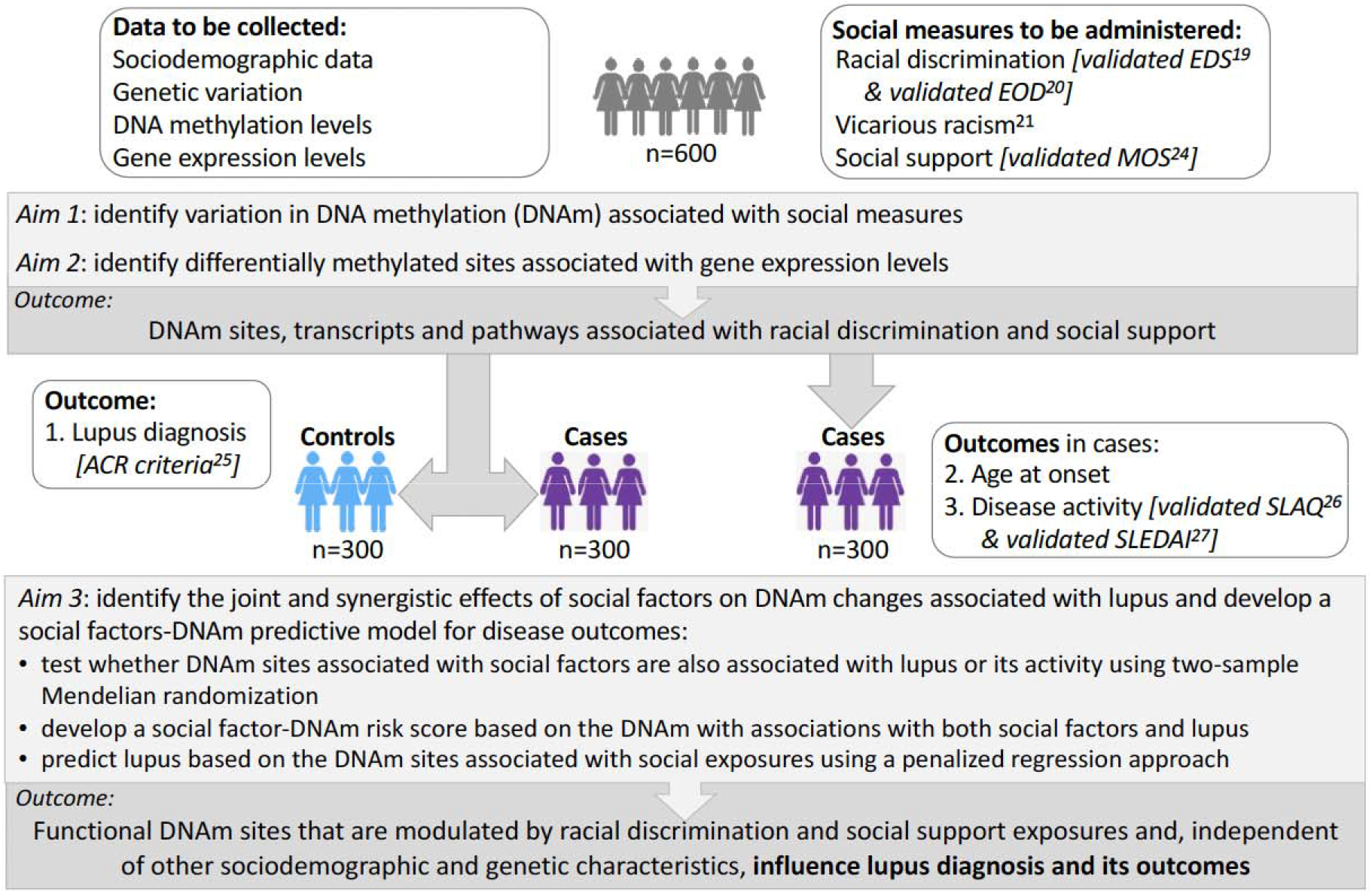
Overall study description. The goal of this study is to identify functional DNAm and/or mediated effects of social factors with effects on the likelihood of lupus and its outcomes.

SELA involves three sites. The Medical University of South Carolina (MUSC) is the coordinating and recruitment center. Wake Forest School of Medicine will generate and lead the statistical and bioinformatics analyses of the genotypic, DNA methylation, and RNA-seq data. Emory University will lead the racial discrimination aspects of this project. A total of 600 participants will be enrolled, 300 cases and 300 controls. Recruitment began in January 2022.

### Study population

SLE disproportionately affects African Americans and women, so it is important to study these health disparity populations. In addition to SLE, social stressors like racial discrimination also place a disproportionate burden on African American women.^17–19^ Since the SLE disparity is already established in African American women, this study focuses on this disparity group, minimizing confounding effects due to ethnicity and gender. Healthy adult controls (age-matched) will be invited to volunteer to respond to questionnaires and donate blood.

Eligibility criteria for patients with SLE and controls are shown in tables 1 and 2. Patients will be primarily recruited from a longitudinal registry of patients for research studies at the Medical University of South Carolina (MUSC).

**Table 1.** Eligibility criteria for cases

| Inclusion criteria | Exclusion criteria |
| --- | --- |
| ○ Self-reported as female<br>○ Age $\geq 18$ years and $\leq 75$ years<br>○ Meet either the 1997 American College of Rheumatology (ACR) revised criteria or the Systemic Lupus Erythematosus International Collaborating Clinics (SLICC) criteria for SLE<br>○ Self-reported as African American or Black<br>○ Able to take part in assessment activities (self-reported questionnaires) | ○ Being a prisoner or institutionalized individual<br>○ Unwilling or unable to give consent<br>○ Development of any infection within 14 days of the initial visit, as reported by the participant<br>○ Current malignancy, as reported by the participant<br>○ Any physical, mental, or other factor that in the judgement of the PI will impact participant safety, privacy, or data integrity |

**Table 2.** Eligibility criteria for controls

| Inclusion criteria | Exclusion criteria |
| --- | --- |
| ○ Self-reported as female<br>○ Age $\geq 18$ and $\leq 75$ years<br>○ No diagnosis of SLE, systemic sclerosis, or other connective tissue disease<br>○ Self-reported as Black or African American | ○ Being a prisoner or institutionalized individual<br>○ Unwilling or unable to give informed consent<br>○ Development of any infection within the past 14 days, as reported by the participant<br>○ Current malignancy as reported by participant<br>○ Any physical, mental, or other factor in the judgement of the PI will impact participant safety, privacy, and data integrity |

All control participants will be recruited via approved advertisements in print and electronic format, including social media platforms. Information about the research study will be provided at educational events for the public. Controls will be self-reported African American females without history of SLE, systemic sclerosis, or other connective tissue diseases that are matched based on patients ‘ age (±5 years).

### Participant Enrollment and Interview

For study enrolment, 300 self-reported African American females with SLE and 300 age-matched African American female controls will be recruited. The informed consent document is sent to potential participants prior to the screening visit, either by mail or email, for their review as requested. Written informed consent of the participant is obtained at the screening visit. Any questions the participant might have will be answered by the study coordinator, physician, or investigator as appropriate, and she will be specifically informed that consent to donate a specimen will in no way obligate her to do so at a future date, nor affect her care in any way. The consent form will allow the participants to consent to future research use and submission of their data to publicly accessible data repositories, hence allowing the sharing of individual, de-identified data. The participants will be informed that their global genetic ancestry estimates will be generated as part of this study and will agree or decline to receive their genetic ancestry composition results after the data are analyzed. If they agree to participate, the study coordinator asks the participant to sign the combined Informed Consent with Health Insurance Portability and Accountability Act (HIPAA) document. No study procedures are performed prior to obtaining written informed consent. Once the participant gives written informed consent, participants will be asked to complete questionnaires and donate biospecimens (see below).

### Questionnaires

The questionnaires will capture sociodemographic and behavioral information (e.g., education, income, occupation, marital status, health insurance, educational attainment, smoking, alcohol use, physical activity, chronic illnesses). Gender identity will be asked (e.g., cis-woman, trans-woman). Since different individuals might prefer more specific terms when defining their race, participants will be asked to describe how they prefer to self-report (e.g., Black-Caribbean, Afro-Latino). They will also be asked if they consider themselves Sea Islanders, and their parents ‘ place of birth.

The questionnaires include two validated measures of racial discrimination, one measure of social support, and one measure of depression (table 3).^18 20–23^ Our primary measure of racial discrimination will be the validated Experiences of Discrimination (EOD) measure.^20^ We will augment this measure with the validated Everyday Discrimination Scale.^23^ We will also perform the same analyses using the measure of vicarious racism stress we previously used; this measure has high internal consistency and reliability (Cronbach α=0.83).^18^ All participants will be asked to respond to healthcare utilization and lost-productivity questionnaire,^24–26^ and to a brief debriefing questionnaire to identify potential distress from responding to the questionnaires. If the participant has a positive screen, we have compiled a list of mental health resources that the study coordinator will be able to provide to the participant. All participants will also receive a short questionnaire to assess their interest in being included in this study ‘s progress and involved in our research.

**Table 3.** Questionnaires used to assess racial discrimination, vicarious racism stress, social support, depression, and SLE disease activity

|  |  |
| --- | --- |
| **Racial discrimination** | Experiences of Discrimination (EOD)<sup>20</sup> |
|  | Everyday Discrimination Scale (EDS)<sup>23</sup> |
| **Vicarious racism stress** | Vicarious racism stress questionnaire from BeWell Study<sup>18</sup> |
| **Social support** | Medical Outcomes Study-Social Support Survey (MOS-SS)<sup>21</sup> |
| **Depression** | Patient Health Questionnaire-8 (PHQ-8)<sup>22</sup> |
| **SLE disease activity** | Systemic Lupus Activity Questionnaire (SLAQ)<sup>26</sup> |
| **SLE disease damage** | Brief Index of Lupus Damage (BILD)<sup>24</sup> |

In addition, all patients with SLE will also complete validated questionnaires that assesses disease activity (SLAQ)^26^ and disease damage (BILD).^24^ The study rheumatologist will evaluate and assess the patients using the Systemic Lupus Erythematosus Disease Activity Index 2000 (SLEDAI-2K).^27^

### Biospecimens

Peripheral blood mononuclear cells (PBMCs), plasma and serum will be isolated from 30 ml of blood collected from participants who agree to participate in this research study. In order to get a current assessment of disease activity (SLEDAI), urine will also be requested from new SLE patients and MUSC patients without recent (within 6 months) visits.

Cell subset composition will be assessed in order to generate reference matrices for the deconvolution of the PBMC methylomic and transcriptomic data. The main PBMC populations (T cells, B cells, NK cells, and monocytes) from each participant will be counted using analytical flow cytometry. Genetic material (DNA and RNA) will be isolated from the PBMCs.

### Data generation and analyses

The Infinium Global Diversity Array (GDA) (Illumina) will be used for genotyping, the MethylationEPIC BeadChip (Illumina) will be used to assess DNA methylation levels, and RNA-seq will be used to measure transcript levels. These assays will be performed at Wake Forest School of Medicine. Genetic variants that pass quality control filters will be imputed to the African Genome Resources Panel. This genotypic data will be used for inference of genetic ancestry and identification of population stratification. The genetic ancestry estimates will be shared with individual participants if requested during the consent process.

The methylation and gene expression data will be deconvoluted in order to adjusting for cell type heterogeneity.^28 29^ Currently, reference datasets for do not match the age, gender, clinical characteristics, or ancestry of the individuals under study.^30^ Of relevance, chronic and acute stressors might also alter blood cell composition, further underscoring the importance of this confounding source of variability when conducting epigenomic and transcriptomic studies using heterogeneous cell mixtures. We will improve the statistical deconvolution by incorporating cell counts from our samples.

To identify DNAm sites associated with life course racial discrimination, we will compute a linear regression model with levels of racial discrimination as a predictor of the methylation level at each CpG site, controlling for age, smoking, white blood cell proportions, SLE status, medication use, and PCA or admixture proportions. Similar regression analysis will be computed to test for an association between DNAm at individual CpG sites and levels of social support. We will also test whether DNAm PhenoAge,^31^ a measure of epigenetic age acceleration, is associated with exposure to racial discrimination, exposure to social support, SLE diagnosis, SLE age-of-onset, and disease activity (as measured by the SLAQ^26^ and SLEDAI^27^ scores).

In order to understand the mechanisms and functional consequences of socially-induced epigenetic changes, we will identify associations between methylation levels and gene expression levels, that is, expression quantitative trait methylation (eQTM) loci. Given the effects of genetic variation on DNA methylation and gene expression, we will also identify associations between genetic variation and methylation levels (cis-meQTL) and associations between genetic variation and gene expression levels (cis-eQTL). Joint effects of racial discrimination and social support on DNA methylation will be modeled through a DNAm regression model that includes social support, racial discrimination, and their interaction, controlling for age, smoking, white blood cell proportions, SLE status, medication use, and PCA or admixture proportions. We will use two-sample Mendelian randomization to identify CpG sites that may mediate the effect of social exposures (racial discrimination and social support) on SLE, including admixture estimates as covariates. We will test if biological pathway-driven DNAm risk scores (e.g., weighted averages of identified DNAm, where weights are the effect size) correlate with SLE or SLE activity (as measured by the SLAQ^26^ and SLEDAI^27^ scores). Finally, we will use machine learning methods (i.e., penalized regression) to develop a social factor-DNAm predictive model for SLE and SLE activity.

### Conceptual Model

Our conceptual model (figure 2) is based on the Biopsychosocial Model, which asserts that African Americans who perceive certain circumstances as racist experience physiological stress responses that can be modulated by adverse or protective sociodemographic (e.g. socioeconomic status) and psychological characteristics (e.g. depression), behavioral factors (e.g. smoking, alcohol use), and coping responses (e.g. ability to mobilize social support) to such experiences.^32^ This conceptual model for African American women integrates social determinants of health with biological factors. Although not a comprehensive model of SLE risk factors, it models how interdependencies among genetic background, environmental exposures, and epigenetic signatures may contribute to increased risk of SLE in African American women. The model (figure 2) depicts how genetic factors (which include ancestry-related variation in allele frequencies and sex) can increase the risk of SLE directly, or through variation in DNA methylation and gene expression. Our analyses will account for this genetic variation by adjusting for genetic ancestry and sex in the model. Since genetic ancestry partly accounts for variation in DNA methylation, it is critical to account for genetic variation among the participants.^9–16^ The model also shows how multiple environmental factors, including social exposures, can directly affect or indirectly affect, through epigenetic dysregulation, risk for SLE. Lupus can also directly affect the response to environmental exposures, causing epigenetic changes. These reverse-causation and confounding factors will be considered during analyses.

**Figure 2.**
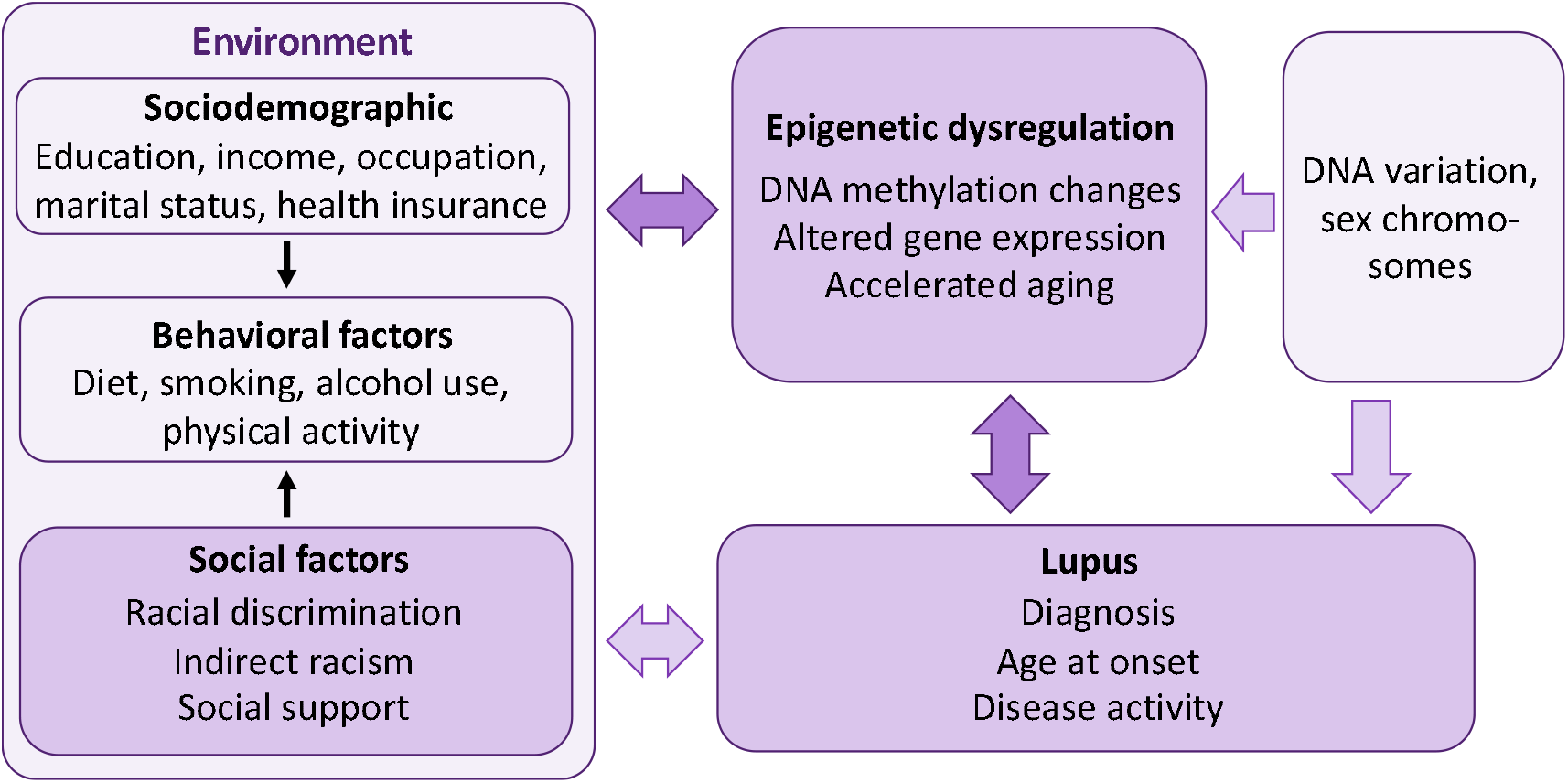
Study description. We posit that social support compensates for the detrimental, independent effect of racism on lupus outcomes, taking into account other sociodemographic and behavioral characteristics, through epigenetic and gene regulatory mechanisms. We will: 1) identify DNA methylation (DNAm) variation associated with exposure to racial discrimination and to social support (measured using validated measures) 2) assess whether these DNAm sites influence gene expression, taking genetic variation into account 3) identify the joint and synergistic effects of social factors on DNAm changes associated with lupus outcomes and 4) develop a social factors-DNAm predictive model for disease outcomes.

### Community engagement and patient involvement

Community engagement is essential to advance understanding of and eliminate racial and ethnic disparities impacting patients with SLE, particularly since economic and social determinants of health, such as poverty, discrimination, and community-level social stressors account for over 40% of the modifiable contributors to healthy outcomes for a population.^33^ Our SLE genetics projects involve African American community members from rural South Carolina and are conducted in cooperation with the Sea Island Families Project (SIFP) Citizen Advisory Committee (CAC).^34^ Interdisciplinary research teams from the MUSC developed community-engaged research projects between academic researchers and Gullah African Americans residing in rural South Carolina, leading to the formation of SIFP-CAC.^34^ This 28-year partnership meets quarterly for sharing of research results and providing guidance and recommendations to new research. The proposed community-engaged study was approved by and will be conducted in cooperation with the SIFP-CAC.

During the study visit, participants will be asked about their willingness to partner with us in research studies trying to understand the causes of autoimmune diseases like SLE. Specifically, they will be asked the following three questions: 1) “Would you like to partner with us, participating with suggestions for and feedback on research studies during annual or bi-annual meetings?”; 2) “If you aren ‘t willing or able to commit to a partnership, are there any research questions or feedback that you have about issues that are important to you, regarding the research we do?”; and 3) “Would you like to be included in annual newsletters reporting the progress of this research study?” Participants who respond affirmatively will be involved as they choose.

## Ethics and Dissemination

This study was approved by the Institutional Review Board at the Medical University of South Carolina (Pro# 112945). Informed consent will be obtained from all participants. All research included in this manuscript conforms with the Declaration of Helsinki.

Progress and results will be presented at national or international conferences, to colleagues at seminars and talks, to the community at patient education events, and the study participants through annual newsletters. Results will be published in peer-reviewed journals in a timely fashion. The investigators will abide by the recent “Updated Guidance on the Reporting of Race and Ethnicity in Medical and Science Journals” published in JAMA.^35^ All final peer-reviewed manuscripts that arise from this proposal will be submitted to the digital archive PubMed Central.

Research data that document, support and validate research findings will be made available at the same time the main findings from the final research data set have been accepted for publication. The investigators will work to facilitate any request made for data produced under this proposal upon publication of data, using standard, university-approved material/data transfer agreements. The genotyping data will be deposited in the National Institutes of Health (NIH) database of Genotypes and Phenotypes (dbGaP), and the DNA methylation and RNA-seq data will be deposited for public access in NCBI ‘s Gene Expression Omnibus (GEO) database. No identifying information will accompany this data. According to the Extramural Institutional Certification of the genotypic data submitted to dbGaP, this data is considered to have particular ‘sensitivities ‘ related to potential for group harm and will only to be made available through controlled-access. In addition to the genomic data, relevant associated data (e.g., phenotype, study protocols) will be concomitantly submitted.

## Discussion

There is increasing awareness and need for integrative mechanistic studies that examine the dynamic interplay of multiple factors across the life course, in order to better understand and address the drivers of health disparities and inform the development of effective interventions. This Social Factors, Epigenomics, and Lupus in African American Women (SELA) study was developed in response to PAR-19-372 (Social Epigenomics Research Focused on Minority Health and Health Disparities), whose purpose is to support epigenomic investigations focused on identifying and characterizing the mechanisms by which social context and experiences, both positive and negative, affect gene function and thereby influence health trajectories or modify disease risk in health disparity populations.

The conceptual framework for this study is based on the socioecological model that emphasizes the importance of integrating societal, community, interpersonal, and individual determinants to understand and address health disparities in SLE.^1^ Social determinants of health span the socioeconomic (employment, income, housing and food security), community (family and social support), neighborhood and physical environment (access to food and housing, crime and violence, safety, transportation, air and water quality), and the health care system (access, quality). Individual determinants include genetic (sex chromosomes, DNA, epigenetic, and gene expression variation) and behavioral factors (diet, smoking, alcohol use, physical and mental health). Since exposures and experiences vary across individuals from different populations, locations, and cultures, it is critical to study population differences in SLE health disparities within the sociocultural context. This is further underscored by both the paucity of disadvantaged communities in research and the genetic and cultural heterogeneity of racial and ethnic groups.

Innovative aspects of this study include the focus on culturally distinct Gullah and non-Gullah African American women, the community partnership, and the integrative analysis of multiple individual and social factors, including risk and protective social effects. This integrative, multiomic research that integrates social and genomic data requires complementary expertise from multiple health centers partnered together to provide expertise in minority health and health disparities, social epidemiology, SLE, genomics, epigenomics, transcriptomics, and statistical methods.

Allowing the participants to further describe how they self-identify beyond the generic “Black or African American” racial category or expand on their gender identities beyond the dichotomous male/female categories, allows for more inclusive participation. Indeed, close to 2% of all individuals are born without being clearly sexed,^36^ and according to the 2020 US Census Bureau “Other” is now the second most common racial group in the US, and the third most common in South Carolina together with “Two or More Races”. ^37 38^ We expect that the increased inclusivity and more granular data might help interpret the results of this study. The option to receive their genetic ancestry estimates has been welcomed by the participants so far.

A potential limitation of this study is the lack of previous research linking specific experiences or behaviors to epigenetic changes in SLE. However, mounting evidence across several traits suggests that epigenetic mechanisms may provide a causal link between social adversity and health disparity.^39^ Another limitation is the current lack of published genome-wide association studies (GWAS) of SLE in patients with African ancestry, thus current polygenic risk score data for African Americans is lacking. If GWAS data in African Americans becomes available during this study, we will integrate the SLE African American polygenic risk scores as potential confounders in our models.

This study will identify the epigenetic mechanisms by which risk and resiliency factors affect gene function and thereby influence SLE in African American women – the most vulnerable and susceptible group to this prototypic autoimmune disease without any safe and effective treatments. The identification of epigenetic mechanisms by which adverse and protective factors affect gene function and thereby influence SLE may inform the development of psychosocial interventions that prevent or mitigate risk exposures, and services or interventions that promote positive exposures. Development of these novel treatments and preventative interventions, as informed by the results of this study, is paramount to the closure of the health disparities gap. Due to the shared etiologic mechanisms, the implications of this research extend across autoimmune diseases and beyond, as an overarching paradigm of the mechanisms for how social stressors physiologically affect human health.

## Data Availability

This protocol has not produced any data.

## Authors’ contributions

EV wrote the original draft preparation; CL and BW contributed with methodology; QQ, LM, QK, IM, SB and LU contributed with investigation; LU, DK, PR contributed with supervision; PR contributed with project administration; CL, BW, TH, GH, SL, EW, DK and PR contributed to the conceptualization of this study and with funding acquisition. All authors reviewed, edited, and agreed on the final version of the manuscript.

## Funding statement

This work is supported by the US National Institute on Minority Health and Health Disparities of the National Institutes of Health (NIH) under Award Number R01 MD015395 (CDL, BJW, TDH, GAH, LQK, IDM, SSL, EMW, DLK, PSR), by the US National Institute of Arthritis and Musculoskeletal and Skin Diseases under Awards Number T32 AR050958 (ELV), P30 AR072582 (BJW, SLB, LAU, EMW, DLK, PSR) and K24 R068406 (DLK), and by the National Center for Chronic Disease Prevention and Health Promotion (NCCDPHP) under Award Number U01 DP006488 (SSL). The content is solely the responsibility of the author and does not necessarily represent the official views of the National Institutes of Health.

## Competing interests statement

The authors declare no conflicts of interest.

## Notes

### Competing Interest Statement

The authors have declared no competing interest.

### Author Declarations

This study was approved by the Institutional Review Board at the Medical University of South Carolina (Pro# 112945).

### Summary of Updates

There was a typo in the originally submitted Figure 1.

